# Quantifying the population-level impact of expanded antibiotic treatment for cholera outbreak management

**DOI:** 10.1101/2024.11.04.24316579

**Authors:** Sharia M. Ahmed, Cormac R. LaPrete, Iza Ciglenecki, Andrew S. Azman, Daniel T. Leung, Lindsay T. Keegan

## Abstract

**Background:** Since 2021, there has been a resurgence in the number of cholera cases, countries affected, and the case fatality risk. Partly due to concerns about antibiotic resistance, current cholera treatment guidelines reserve antibiotics for severely symptomatic cases, recommending only supportive care (e.g., oral rehydration) for non-severe cases. However, it has been suggested that the reduction in transmissibility from antibiotic treatment may result in circumstances under which treating mild or moderate cases with antibiotics may have population-level benefits. We developed a compartmental model of cholera transmission in a non-endemic setting to quantify the potential impact of expanded antibiotic treatment on disease burden and antibiotic use. Through simulations, we evaluated different outbreak scenarios, by varying the reproductive number, care-seeking behavior, and proportion of non-severe cases receiving antibiotics. We found that expanding antibiotic treatment could significantly reduce the final outbreak size under certain outbreak characteristics. Under these different transmission scenarios, treating non-severe symptomatic infections with antibiotics decreased cholera transmission and, in some cases, the total number of antibiotic doses used. In high transmission settings, the benefits of expanded treatment are less pronounced and the strategy may lead to increased antibiotic use, potentially increasing the risk of antibiotic resistance. We show that the effectiveness of expanded antibiotic treatment is highly dependent on achieving high care-seeking rates among non-severely symptomatic infections and tailoring the approach to specific outbreak conditions. While expanding antibiotic eligibility could enhance outbreak control in some settings, careful consideration of antibiotic resistance risks is necessary in high-transmission contexts.

## Introduction

Cholera, caused by the toxigenic bacterium *Vibrio cholerae* O1/O139, remains a significant public health threat [1]. It is characterized by severe acute watery diarrhea that can cause death within hours if left untreated [1,2]. Despite its severe clinical manifestations, the true burden of cholera is not well characterized, as only approximately 10% of cases experience severe symptoms and it has been estimated that the vast majority of infections are asymptomatic or unreported [3–6]. A 2015 global burden analysis estimated 2.86 million cases (uncertainty range 1.3–4 million cases) and 95,000 deaths (uncertainty range 21,000–143,000 deaths) in endemic countries annually [7]. Although the global burden of cholera has been declining in recent decades, a notable resurgence in cases, countries affected, and the case fatality rate has been described since 2021 [8].

Patterns of cholera transmission vary between endemic regions and non-endemic regions, in part due to protective immunity [9]. Importantly, the estimated durability of natural immunity varies from several months to 10 years [10]. Subclinical cholera infections may confer lower protection than clinical infections [11]. As a result, in endemic regions where local transmission has been detected over the past 3 years, incidence of cholera is highest among young children, as young children are the least likely to have previous exposure and therefore immunity. Whereas in non-endemic (outbreak) settings where cholera does not regularly occur, attack rates among children and adults are similar, as the entire population has a similarly low or non-existent level of pre-existing immunity [12,13].

Cholera is typically an easily treatable disease, with rehydration serving as the cornerstone of treatment. Non-severe cholera cases can be successfully managed with oral rehydration solution and severe cases with intravenous rehydration. When administered promptly, rehydration therapy can reduce the case fatality risk to below 1% [1,6,14,15].

Antibiotics, though available for cholera treatment, are generally reserved for severely dehydrated patients or those with high-risk conditions such as pregnancy or severe acute malnutrition, in part due to concerns about antibiotic resistance [14–16]. While antibiotics reduce the duration of symptoms and stool volume, their use is not typically recommended for non-severely dehydrated cases as the benefit to the patient is modest relative to the risk of development of antibiotic resistance at the population level [15,17,18]. Antibiotics do however confer benefits related to transmissibility: infections untreated with antibiotics shed *V. cholerae* for up to 10 days after symptom resolution, contributing to community transmission [1]. In contrast, antibiotic treatment reduces the duration of bacterial shedding by up to 90% and likely the concentration of infectious bacteria in the stool, thereby considerably reducing cases’ transmission potential [15,16].

Currently, treatment guidelines reserve antibiotics for the most severe infections in part due to the modest benefit conferred to non-severely symptomatic infections and partly due to concerns about antibiotic resistance [15]. When indicated, the recommended treatment for cholera is a single dose of doxycycline, though azithromycin and ciprofloxacin can be used in certain situations [15]. Antibiotics are often under-regulated in areas where cholera is most likely to occur, and people frequently self-medicate with antibiotics before presenting to formal clinical care [19–21]. These self-medicated antibiotics are often inappropriate to treat cholera and further contribute to the development of antibiotic resistance [19]. Recently, it has been proposed that the reduction in transmissibility associated with antibiotic treatment could offer public health benefits by curtailing outbreak transmission [22]. Expanding antibiotic treatment guidelines to include mild and moderately symptomatic infections may result in fewer antibiotic doses used over the course of an outbreak by reducing transmission, resulting in fewer infections compared to current treatment guidelines. In this study, we develop and apply a compartmental model of cholera transmission to quantify the impact of expanding antibiotic treatment to include mild or moderately symptomatic infections. Our objective was to assess how treatment of those with moderate/some dehydration could impact the overall burden of cholera in an outbreak as well as the risk of developing antibiotic resistance.

## Methods

### Model

To assess the impact of expanded antibiotic treatment guidelines on cholera outbreaks, we developed and analyzed a compartmental model of cholera transmission in a non-endemic setting (Figure 1). Susceptible (*S*) individuals, with no immunity from prior infection or vaccination become exposed (*E*) upon successful transmission and then progress to one of the infectious compartments (*I*). We differentiate within the infectious compartments based on symptomatology: asymptomatic (*I*_*A*_), non-severely symptomatic infections, including both mild and moderately symptomatic (*I*_*M*_), (hereafter referred to collectively as “moderate”), and severely symptomatic (*I*_*S*_) as defined by dehydration. Additionally, we differentiate within the non-severely symptomatic infections based on care seeking behavior: seeking care (*I*_*U*_) or not seeking care (*I*_%_).

**Figure 1:**
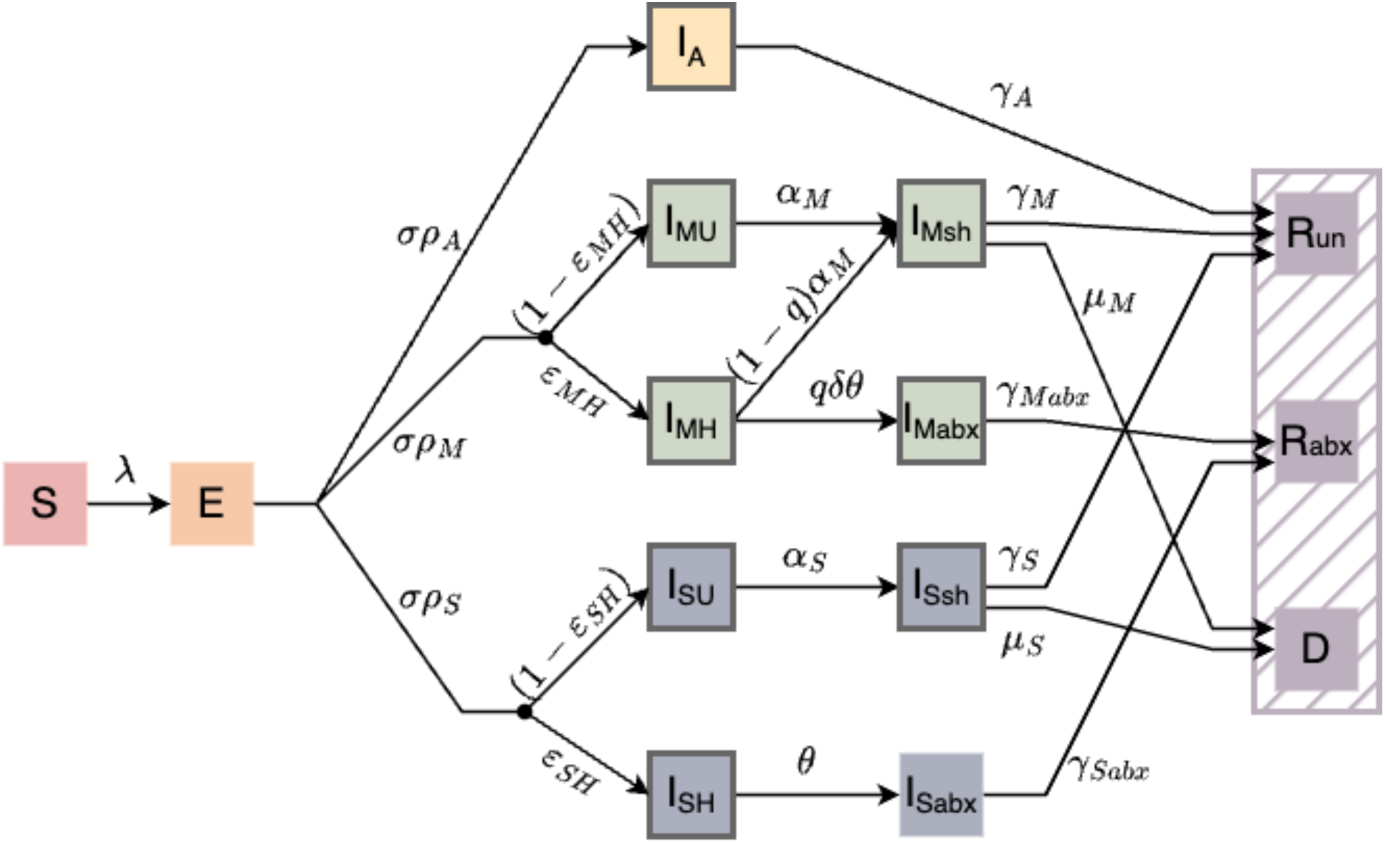
Compartmental diagram of the cholera transmission model. All individuals start as susceptible (*S*) and become exposed (*E*) at a rate λ. Exposed individuals transition to the infected (*I*) compartment and we differentiate by symptoms (*I*_*A*_, *I*_*M*_, *I*_*S*_, asymptomatic, non-severely, or severely symptomatic, respectively) and by care seeking behavior (*I*_*H*_,*I*_T,_, not care seeking and care seeking, respectively). All severely symptomatic infections are treated with antibiotics whereas not all non-severely symptomatic infections who seek treatment receive antibiotics. The proportion of healthcare seeking non-severely symptomatic infections who receive antibiotics is governed by *q*. Untreated infections, both non-severe and severe, continue to shed for a longer duration following the resolution of symptoms (*I*_*Msh*_, *I*_*Ssh*_), occurring at rate *α*_*M*_, *α*_*S*_, respectively. Non-severely symptomatic infections who are treated with antibiotics continue to shed for a shorter duration following treatment (*I*_*Mabx*_), occurring at rate *δθ*, whereas severely symptomatic infections who are treated with antibiotics remain in a treatment facility and do not contribute to transmission (*I*_*sabx*_). Compartments with a dark grey outline indicate they contribute to transmission. Infectious individuals either recover (*R*, at rates *γ*_*A*_, *γ*_*M*_, *γ*_*Mabx*_, *γ*_*S*_, *γ*_*Sabx*_) or die (*D*, at rates *μ*_*M*_, *μ*_*S*_) and we differentiate between individuals who recover without antibiotic treatment (*R*_*un*_) and those who recover with antibiotics (*R*_*abx*_) to compare the number of doses used under different treatment scenarios.

We do not attempt to parameterize care seeking behavior, rather, we vary the proportion of non-severely symptomatic infections who seek care between simulations. Due to the nature of their symptoms, we assume that all severely symptomatic infections who seek care (*I*_*SH*_) receive antibiotic treatment (*I*_*Sabx*_) and that severely symptomatic infections are much more likely to seek treatment. We model the effect of varying the proportion of non- severely symptomatic infections that seek (*I*_*MH*_) and receive antibiotic treatment (*I*_*Mabx*_).

We assume that asymptomatic infections never seek treatment and thus do not further divide this compartment by treatment.

Since infected individuals untreated with antibiotics continue to shed *V. cholerae* for up to 10 days post-symptom resolution [23], we assume that they continue to contribute to transmission after the conclusion of symptoms, but at a reduced rate (*I*_*Msh*_, *I*_*Ssh*_, for non- severely and severely symptomatic infections, respectively). For non-severely symptomatic infections treated with antibiotics (*I*_*Mabx*_), we assume that they contribute to transmission after receiving antibiotic treatment but at a reduced rate and for a reduced duration. In contrast, we assume that severely symptomatic infections that seek care (*I*_*Sabx*_) are admitted to a treatment center and therefore no longer shed into the community, and thus do not contribute to transmission after receiving antibiotic treatment. In general, we parameterize our model to ensure key rates for non-severely symptomatic infections, such as treatment seeking (θ) and proportion in each symptom class (*ρ*_*A*_, *ρ*_*M*_, *ρ*_*S*_) are proportional to those for severe infections. For full model equations, *see supplemental material*.

### Scenarios

We evaluate the impact of treating mild and moderately (non-severe) symptomatic infections with antibiotics over different outbreak scenarios. These scenarios are characterized by varying the effective reproductive number, the proportion care seeking, and proportion treated with appropriate antibiotics.

Because cholera outbreaks have high variability in the reported effective reproductive number by outbreak setting, we designed three scenarios based on these different transmission characteristics at the start of the outbreak: low *R*_*e*_ = 1.3 − 1.5 and intermediate *R*_*e*_ = 1.6 − 2.0 based off outbreaks from Africa [26–30] and high *R*_*e*_ = 2.3 −2.8 based off outbreaks from the Americas [24,25,31,32].

Our primary interest lies in understanding the impact of expanding antibiotic treatment eligibility guidelines to include non-severely symptomatic infections. However, the proportion of non-severely symptomatic infections that receive appropriate antibiotics is governed both by the proportion that seek care (*ε*_*MH*_) as well as the proportion of care-seeking non-severely symptomatic infections who are treated (*q*). Care-seeking behavior among non-severely symptomatic infections is not well characterized and is likely influenced by the current treatment guidelines that limit antibiotics to severely symptomatic infections. As such, we explore five scenarios for care seeking: 5%, 25%, 50%, 75%, and 100% of non-severely symptomatic infections seeking care (five values of *ε*_*MH*_).

Within the scenarios described above, we also vary the proportion of non-severely symptomatic infections who have sought care that receive antibiotic treatment (“proportion treated with antibiotics”, derived from *q*, see supplement) from no one (current treatment guidelines) to everyone (all care-seeking non-severely symptomatic infections receive antibiotics).

### Simulations

We simulate our model using R statistical software [45] over the range of parameter values presented in Table 1 assuming a total population of 4 million, based on average cholera outbreak sizes[46,47]. Since we are modeling a non-endemic outbreak, we initialize our model with 10 exposed individuals and all others susceptible. We simulate each scenario 1000 times until stochastic extinction.

**Table 1:** Summary of the model parameters (and available sources) used in model.

| Parameter | Meaning | Value/Range | Source |
| --- | --- | --- | --- |
| $R_e$ | Effective reproductive number | Low: 1.3 – 1.5<br>Intermediate: 1.5–2.0<br>High: 2.3 – 2.8 | [2,24–32] |
| $1/\sigma$ | Latent period | 1.3-1.6 days | [33] |
| $\rho_A, \rho_M, \rho_S$ | Proportion of infected individuals in each symptom class such that:<br>$\rho_A + \rho_M + \rho_S = 1$ | Asymptomatic: 0.65–0.85<br>(see *supplement for additional ranges*)<br>Non-severe: 0.09 – 0.28<br>Severe: 0.05 –0.1 | [34–37] |
| $\varepsilon_{MH}, \varepsilon_{SH}$ | Proportion of non-severe or severely symptomatic infected individuals who seek care, respectively (proportion care-seeking) | Non-severe: 0–1, varies based on scenario<br>Severe: 0.5 –0.9 | [37,38] |
| $\alpha_M, \alpha_S$ | Time to developing symptoms for untreated infections (in days) | Non-severe: 2.5-5 days<br>Severe: 3.34-10 days | [39,40] |
| $1/\theta$ | Time to treatment for severe infections (in days) | 0.25-1.5 days | Varies based on patient behavior |
| $1/\delta$ | Relative increase in time to treatment for non-severe relative to severe infections | 2 | Based on patient behavior |
| $q$ | Non-severely symptomatic infection treatment effort (see *supplement for additional details*) | Varies by scenario | |
| $1/\gamma_A, 1/\gamma_M, 1/\gamma_S$ | Time to recovery without antibiotic treatment (in days) | Asymptomatic: 3.4-10 days<br>Non-severe: 4.2-10 days<br>Severe: 5-10 days | [39–44] |
| $1/\gamma_{M_{abx}}, 1/\gamma_{S_{abx}}$ | Time to recovery with antibiotic treatment (in days) | Non-severe: 1.67-3.34 days<br>Severe: 1 day | [39–44] |
| $1/\mu_M, 1/\mu_S$ | Time to death (in days) | Asymptomatic: do not die<br>Non-severe: 1-2 days<br>Severe: 0.5 –1 day | [39–41,44] |

Many key parameters are not well characterized for cholera. As such, we use Latin hypercube sampling (LHS) to sample across the full parameter uncertainty range [48,49]. LHS ensures comprehensive exploration of the parameter space by sampling each interval only once, allowing for the distinction between variability in outcomes driven by the stochastic nature of the model from that arising due to parameter uncertainty [49]. In other words, if our range of parameter value encompasses the true parameter value, the outcome metric should also encompass the truth. We constrain all parameters to ensure that no draws from the LHS yield epidemiologically implausible values.

### Epidemiologic Outcomes

We evaluate the scenarios across several metrics, including the total number of infections (final size), the total number of antibiotic doses used over the course of the outbreak, the number of infections under expanded treatment minus the number of infections under current treatment guidelines (infections averted), as well as the number of doses used under expanded treatment minus the number of doses used under current treatment guidelines (additional doses used).

## Results

Through simulation, we show that expanding antibiotic treatment guidelines to include non-severely symptomatic infections can substantially reduce the burden of cholera in low and intermediate transmission settings, especially when rates of care-seeking behavior is high. In high transmission settings, the impact of expanded antibiotic treatment guidelines is less pronounced but still offers marginal reductions in the final outbreak size.

We find that treating non-severely symptomatic infections with antibiotics decreases the final size of the outbreak across all care seeking scenarios and transmission scenarios except in high *R*_*e*_ outbreaks with very low proportion of care seeking (5%) among non-severely symptomatic infections (Figure 2). While treating non-severely symptomatic infections with antibiotics almost always decreases the final outbreak size, the effect is largest when more non-severely symptomatic infections seek care and receive appropriate antibiotics as well as when the effective reproductive number is low.

**Figure 2:**
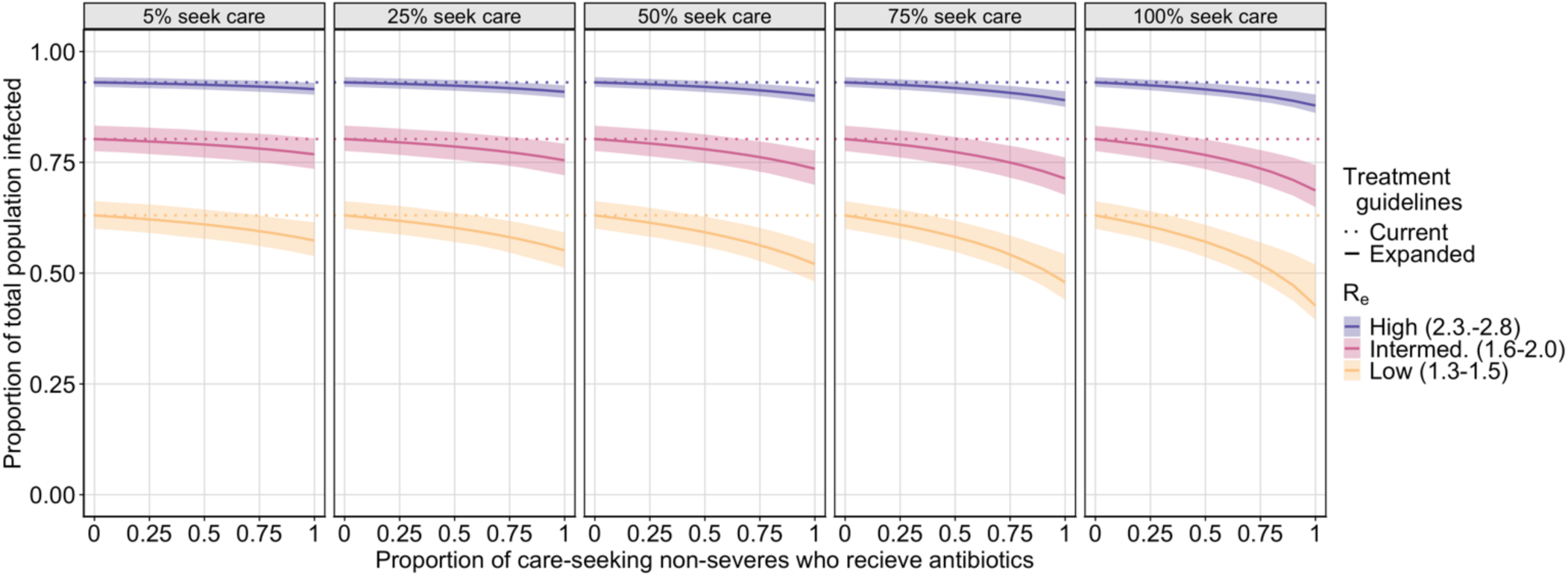
P**l**ot **of the final outbreak size by the proportion of care-seeking non-severely symptomatic infections treated with antibiotics.** Each plot shows the final proportion of the population infected by the proportion of care-seeking non-severely symptomatic infections who receive treatment for low (yellow), intermediate (pink), and high (purple) effective reproductive numbers, for a different percent of non-severely symptomatic infections who seek treatment (5%, 25%, 50%, 75%, 100%). The solid line indicates the mean estimate under expanded treatment guidelines, the shaded region represents the 25% and 75% quantiles, and the dashed line shows the final size of the outbreak under current antibiotic treatment guidelines (treating no non-severely symptomatic infections).

While treating non-severely symptomatic infections with antibiotics can substantially reduce cholera transmission, we show that expanded antibiotic eligibility alone is not sufficient to significantly reduce transmission unless it is coupled with high care-seeking rates.

The development of antibiotic resistance remains a significant concern that informs current cholera treatment guidelines. Although we did not explicitly model the evolution of resistance, we use the number of antibiotic doses administered as a proxy for selective pressure. As such, we evaluated our scenarios based on the number of antibiotic doses used and categorized them by the scale of impact: isolated, expanding, and broad. Our simulations reveal three distinct regions within the range of tested parameters (Figure 3). In one region, extending antibiotic treatment guidelines to include non-severely symptomatic infections confer only isolated benefits, as using additional doses does not reduce transmission sufficiently to avert more than one infection over the course of the outbreak (yellow points, Figure 3). The next region is such that extending antibiotic treatment criteria provides an expanding benefit, whereby treating non-severely symptomatic infections with antibiotics reduces transmission sufficiently such that each additional dose averts more than one infection (green points, Figure 3). In the final region extending antibiotic treatment criteria results in broad benefits where fewer antibiotic doses are used over the course of an outbreak compared to current treatment practices (purple points, Figure 3). Which region a simulation falls into is governed by both *R*_*e*_ and the proportion of non-severely symptomatic infections treated with antibiotics. As *R*_*e*_ increases, the benefits of treating non-severely symptomatic infections with antibiotics shifts from population-level benefits and antibiotic dose reduction to primarily individual-level benefits. Likewise, as the proportion of non-severely symptomatic infections treated with antibiotics increases, the population-level benefit of expanded antibiotic use becomes more pronounced.

**Figure 3:**
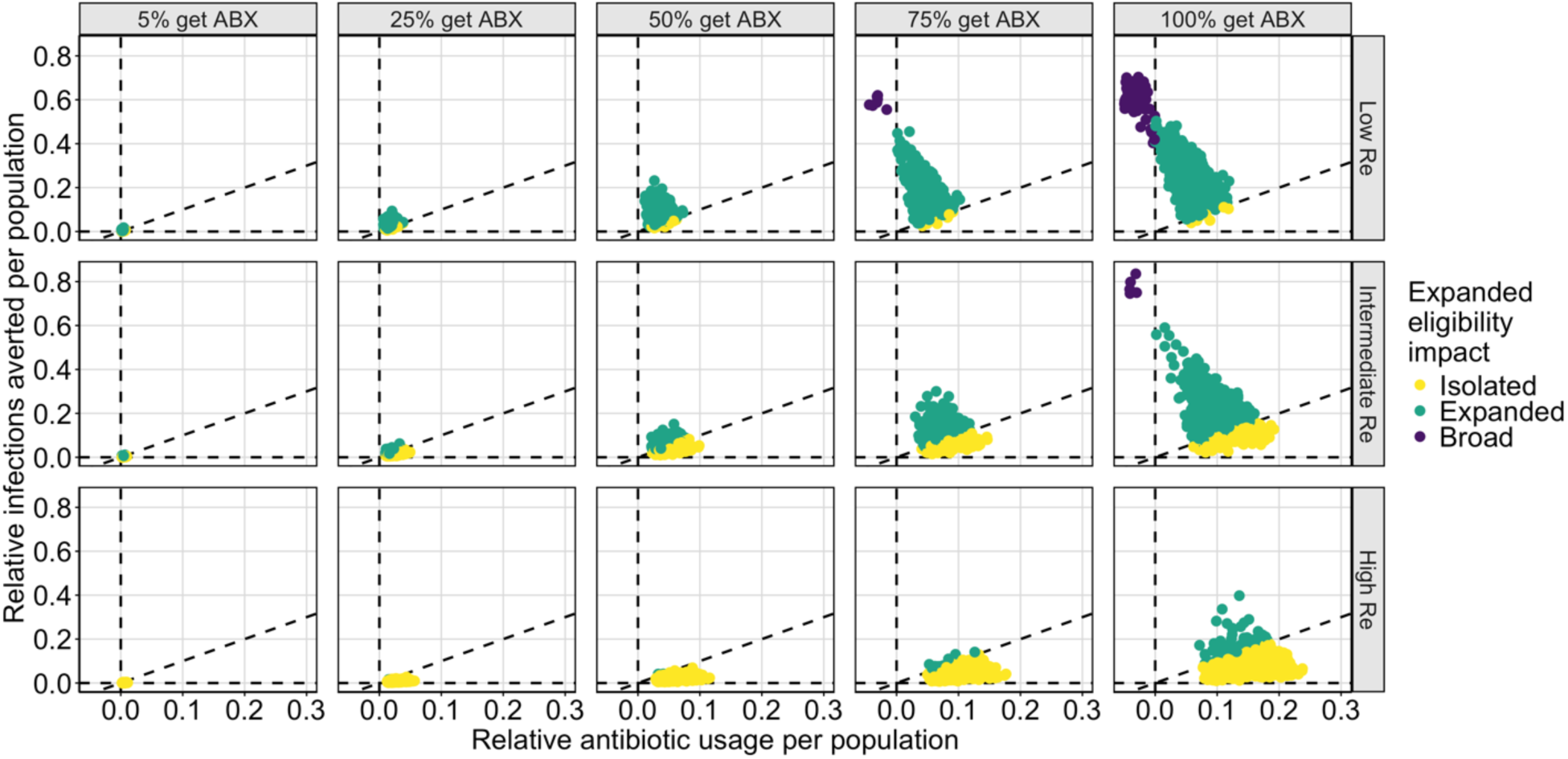
P**l**ot **of the population-level impact of expanded antibiotic treatment guidelines.** Each plot compares the relative infections averted per population to the relative antibiotic usage per population. The relative reduction in infections is calculated by the ratio of the number of infections in each expanded antibiotic treatment scenario presented compared to the simulation of current antibiotic treatment guidelines (treating no non-severely symptomatic infections with antibiotics) using the same LHS sampled parameters, normalized by population size. Similarly, the relative antibiotic usage is calculated by the ratio of antibiotic doses used in each scenario presented compared to the simulation of current antibiotic treatment guidelines (treating no non-severely symptomatic infections with antibiotics) using the same LHS sampled parameters, normalized by population size. Each plot represents a different proportion of non-severely symptomatic infections seeking care who receive antibiotic treatment (5%, 25%, 50%, 75%, 100%) and a different *R*_*e*_ scenario (low (*R*_*e*_ = 1.3 − 1.5), intermediate (*R*_*e*_ = 1.6 − 2.0), high (*R*_*e*_ = 2.3 − 2.8)). The outcomes are split into three regions by the impact of expanded eligibility criteria: between the dashed line along the x-axis (expanding criteria averts no infections) and the diagonal dashed line (each addition dose of antibiotics deployed averts one infection), yellow points represent simulations in which expanded eligibility only has isolated benefits; between the diagonal dashed line and the vertical dashed line (no additional doses are used to avert infections), green points represent simulations in which expanded eligibility results in each dose preventing more than one additional infection; and in the region left of the vertical dashed line, purple points represent simulations in which expanded eligibility results in fewer doses used over the course of the outbreak than compared to current antibiotic treatment guidelines.

To further explore the impact of expanded treatment on the number of antibiotic doses used over the course of the outbreak, we evaluate how the proportion of care-seeking non-severe individuals who receive appropriate antibiotics impacts the proportion of the total population who receive appropriate antibiotics (Figure 4). As with infections averted, when the transmission rate is lower and when more non-severely symptomatic infections are treated, treating non-severely symptomatic infections with antibiotics has greater public health benefits. Indeed, when *R*_*e*_ is low, increasing the proportion of non-severely symptomatic infections treated with antibiotics results in fewer antibiotic doses used. For intermediate and high transmission settings, increasing the proportion of non-severely symptomatic infections treated with antibiotics only results in an increase in the proportion of the total population treated with antibiotics.

**Figure 4:**
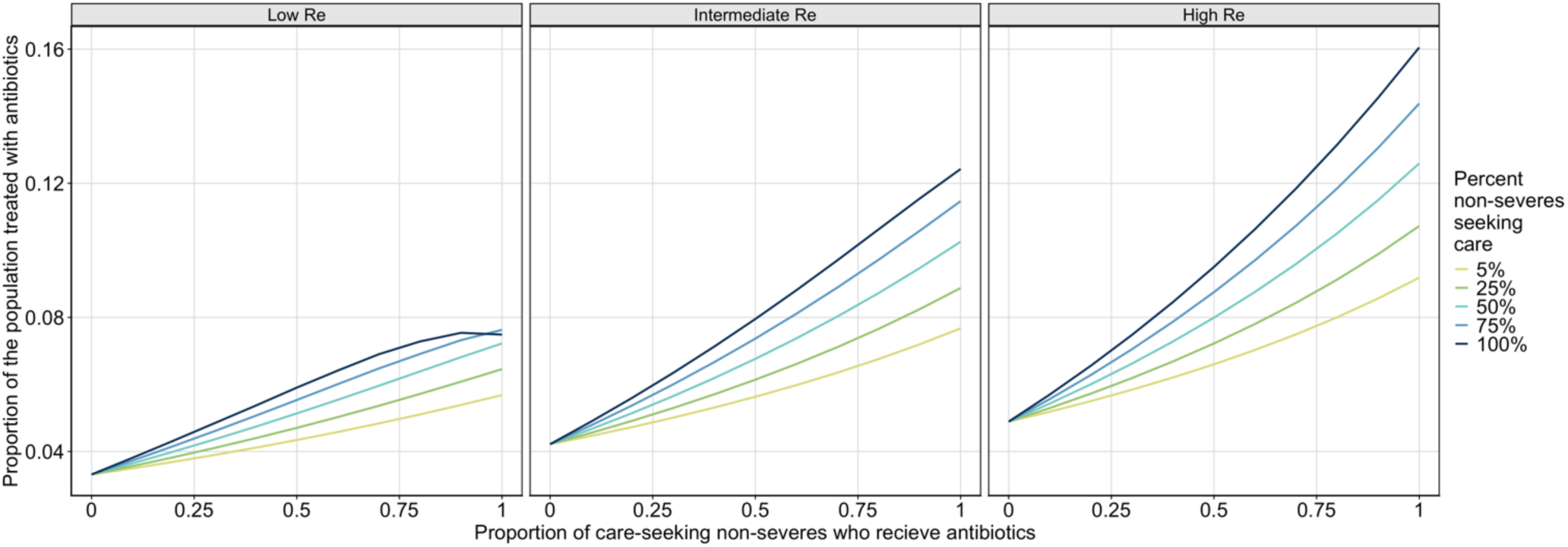
P**l**ot **of population-level antibiotic treatment.** Each plot shows the proportion of the total population who receive appropriate antibiotics, by the proportion of care-seeking non-severely symptomatic infections that receive appropriate antibiotics, separated by *R*_*e*_scenario (low (*R*_*e*_ = 1.3 − 1.5), intermediate (*R*_*e*_ = 1.6 − 2.0), high (*R*_*e*_ = 2.3 − 2.8)). For each *R*_*e*_ scenario, we show five values for the proportion of non-severely symptomatic infections that seek care, 5% (yellow), 25% (green), 50% (teal), 75% (blue), 100% (navy), and vary the proportion of care-seeking non-severely symptomatic infections who receive antibiotics from 0 – 100%.

## Discussion

This study builds on prior research suggesting that, under certain circumstances, expanding antibiotic treatment of cholera cases can provide population-level benefits by reducing transmission and limiting an outbreak size such that fewer antibiotic doses are used over the course of the outbreak. While previous research proposed a theoretical mechanism, our work explores the specific conditions where these benefits can be achieved for cholera. Our findings show how treating non-severe cholera cases with antibiotics can help control outbreaks and highlight the critical role of linking these cases to healthcare in achieving impact. We detail how outbreak specific quantities such as the effective reproductive number and the proportion of non-severely symptomatic infections that seek care impact the effectiveness of this strategy.

The effectiveness of expanding antibiotic treatment criteria improves as the proportion of non-severely symptomatic infections seeking care increases. In the majority of outbreaks with low to intermediate transmission rates (measured by the reproductive number), such as those seen in most cholera outbreaks [21–25], reducing transmission can substantially lower the proportion of the population infected, supporting the theoretical findings that expanded antibiotic access can enhance outbreak control.

A key distinction emerges between low and high *R*_*e*_ settings. In lower *R*_*e*_settings, treating a high proportion of non-severely symptomatic infections reduces both cholera burden and number of antibiotic doses used over the course of the outbreak. However, as *R*_*e*_ increases, these benefits disappear, though an intermediate range exists where each additional dose of antibiotics used averts more than one infection, still providing a public health benefit. In high *R*_*e*_settings, treating non-severely symptomatic infections may reduce the outbreak size, but it requires treating a larger portion of the population. These results suggest that treating non-severe cholera cases may be effective in low *R*_*e*_ settings but may carry a risk of antibiotic resistance in high-transmission contexts, underscoring the importance of tailoring treatment strategies to the specific outbreak context. In this paper, the only intervention we consider is expanding antibiotic treatment guidelines, however this could be combined with other interventions as a component in a large outbreak containment strategy.

Across all simulations, the most important factor governing the effectiveness of treating non-severely symptomatic infections with antibiotics is achieving high antibiotic treatment rates. Often the focus on clinical care during outbreaks is at cholera treatment centers, typically located at secondary and tertiary health structures. While there are gaps in our understanding of care seeking behavior for non-severely symptomatic infections, experience in endemic settings suggest that most people with non-severe disease will seek care at pharmacies or lower-level health facilities, frequently obtaining inappropriate or non-indicated antibiotics [20,21,50–52]. Developing strategies to expand access to appropriate antibiotics could have a critical impact on achieving sufficiently high coverage of antibiotics among non-severe cases to reduce cholera transmission while also improving antibiotic stewardship. Expanded access could be coupled with the use of cholera rapid diagnostic tests, which are typically not used for clinical decision making, to improve the specificity of targeting people with diarrhea caused by *Vibrio cholerae* O1.

Addressing barriers to care seeking behavior is necessary to achieve sufficiently high treatment rates. Cholera case-area targeted interventions are an active and promising area of research and may offer a strategy to achieve these desired antibiotic treatment rates [53–55].

Some of the limitations of our study include that many key model parameters are not well characterized for cholera. Accurate estimates of, in particular, *R*_*e*_ and the proportion of non-severely symptomatic infections seeking treatment are crucial. Our model only considers non-endemic settings, assuming no prior immunity. Further analysis is needed to assess how prior immunity or vaccination affects our results. While we do not model immunity directly, in a supplementary analysis, we evaluate the impact of varying the ratio of asymptomatic to symptomatic infections, one of the expected main differences between an outbreak setting to an endemic setting. We find that our final size results are robust to variations in the proportion asymptomatic, but for outbreaks with a very high proportion of infections that are asymptomatic (consistent with highly endemic settings), the broad impacts from expanding antibiotic eligibility disappear (Figure SI1). Conversely, for outbreaks with a very low proportion of infections that are asymptomatic (consistent with totally naïve settings), not only are broad and expanded impacts more pronounced (Figure SI2), in low *R*_*e*_ outbreak settings, expanding antibiotic treatment guidelines has such a substantial benefit that it halts transmission so effectively that it can prevent an outbreak from ever taking off (Figure SI3). Thus, we expect expanded treatment guidelines may have a smaller impact in endemic settings than outbreak settings.

Additionally, we only consider three symptom categories – severely symptomatic, non-severely symptomatic, and asymptomatic infections. A more granular distinction within the non-severely symptomatic compartment may refine treatment estimates. Since infectiousness correlates with symptom severity [5], it may suffice to target the most symptomatic non-severely symptomatic infections. Achieving a high enough treatment coverage may be a barrier to utilizing this strategy as an outbreak containment strategy, particularly in resource-limited settings where both antibiotics and healthcare infrastructure may be constrained.

Our study builds on phenomenological findings which suggest that treating non-severely symptomatic infections can reduce the number of antibiotic doses needed and may therefore lower the selective pressure for cholera to develop resistance. However, uncertainties in parameter estimates raise concerns about balancing these benefits with the risks of promoting antimicrobial resistance. Future studies aimed at improving characterization of key parameters or a clinical trial aimed at validating these model results and assessing the feasibility of implementing expanded treatment guidelines are a critical next step towards implementing these findings. Careful monitoring of key parameters and resistance patterns will be crucial in ensuring that the benefits of expanded antibiotic treatment criteria do not increase the risk of developing antibiotic resistant cholera.

## Data Availability

This manuscript only uses publicly available data.

## Acknowledgements

The authors thank Kate Alberti for helpful discussions regarding framing the research project.

## Supplemental Information

### Methods

#### Model Equations

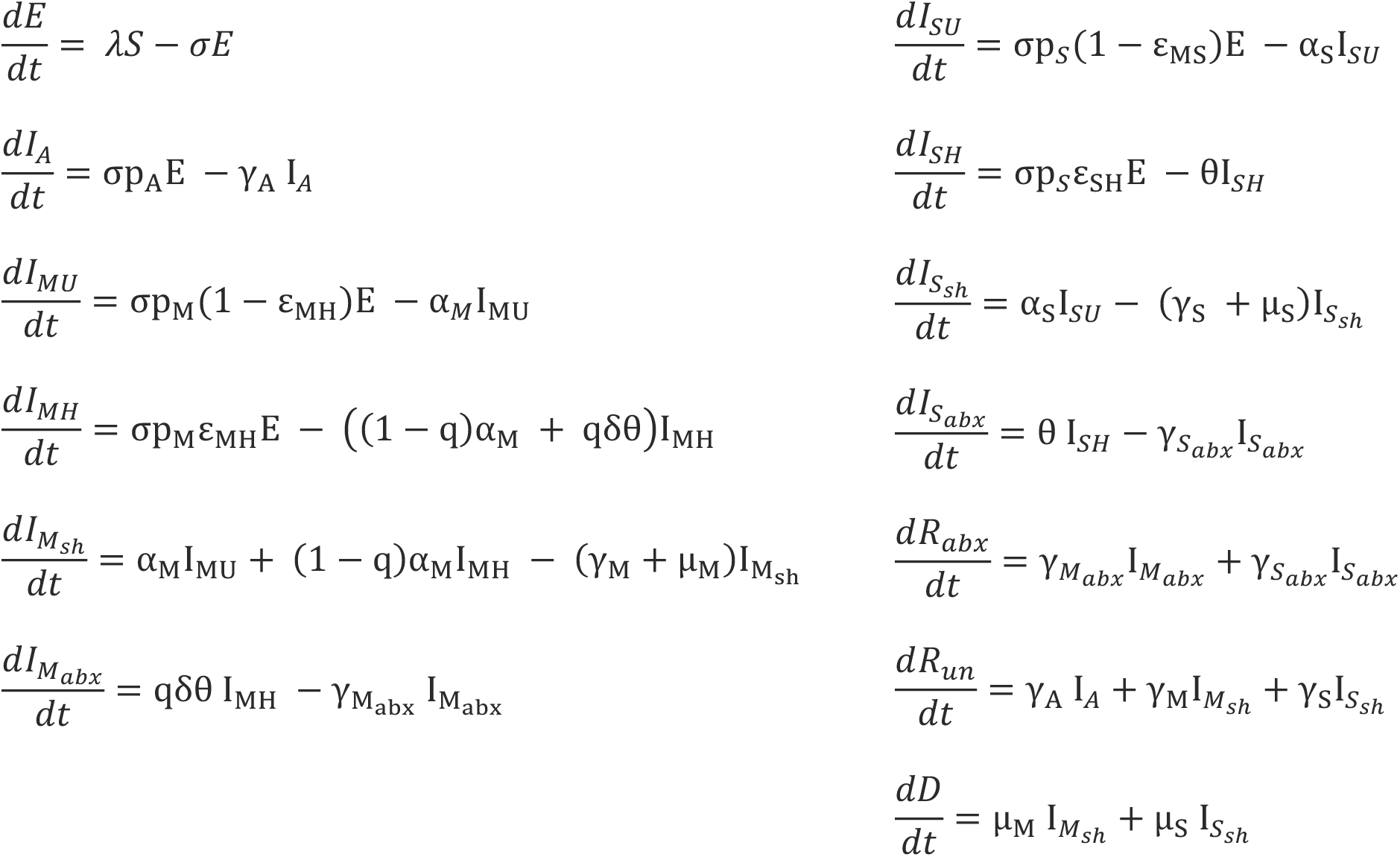

Proportion of care-seeking non-severely symptomatic infections that receive antibiotics

As derived from LaPrete et al. [22], the proportion of care-seeking non-severely symptomatic infections that receive antibiotics, *M*_*abx*_, is given by:

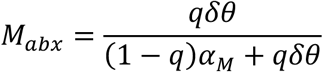

**Table S1:** Summary of the transmission parameters (and available sources) used in model.

| Parameter | Meaning | Value | Source |
| --- | --- | --- | --- |
| $\nu_A$ | Reduction in infectiousness, relative to untreated severe infections, for being asymptomatic | 0.15–0.35 | Expert elicitation |
| $\nu_M$ | Reduction in infectiousness, relative to untreated severe infections, for being non-severely symptomatic | 0.45–0.75 | Expert elicitation |
| $\nu_{sh}$ | Reduction in infectiousness, relative to untreated severe infections, for no longer being symptomatic but still shedding | 0.3–0.6 | [39–41] |
| $\nu_{abx}$ | Reduction in infectiousness, relative to untreated severe infections, for having received antibiotics but still shedding | 0.3–0.7 | Expert elicitation |

*Assessment of the ratio of asymptomatic to symptomatic infections:* Although our study aims to explore non-endemic settings, since the ratio of asymptomatic to symptomatic infections varies by location, we conducted an additional sensitivity analysis to quantify the impact of the ratio of asymptomatic to symptomatic infections on disease burden and number of antibiotic doses used under expanded antibiotic treatment guidelines. Our results in the main text show asymptomatic infections representing 65%-85% of all infections. Here we explore increasing this ratio to 90%-99%, as was reported in Bangladesh[56] and lowering it to 17%-33%, as was found in Haiti [57].

## Results

In a supplementary analysis, we vary the ratio of asymptomatic to symptomatic infections from 65%-85% up to 90%-99%, as well as down to 17%-33%. Overall, we find that our final size results are robust to changes in the ratio of asymptomatic to symptomatic infections and that treating non-severely symptomatic infections reduces the final size regardless of this ratio. However, we find that the broad impacts from expanded eligibility disappear for very high proportion asymptomatic (90%-99%) (**Figure SI1**). Simultaneously, for very high proportion of asymptomatic, we find that the total proportion of the population receiving antibiotics is very small, since most infections are asymptomatic and therefore do not seek treatment. Conversely, when asymptomatic infections represent a smaller proportion of the population (17%-33%), the proportion of simulations with expanded or broad impacts from expanded eligibility become more pronounced (**Figure SI2**). Indeed, for some simulations, the benefit is so substantial that it can halt the outbreak before it can take off (**Figure SI3**).

**Figure SI1:**
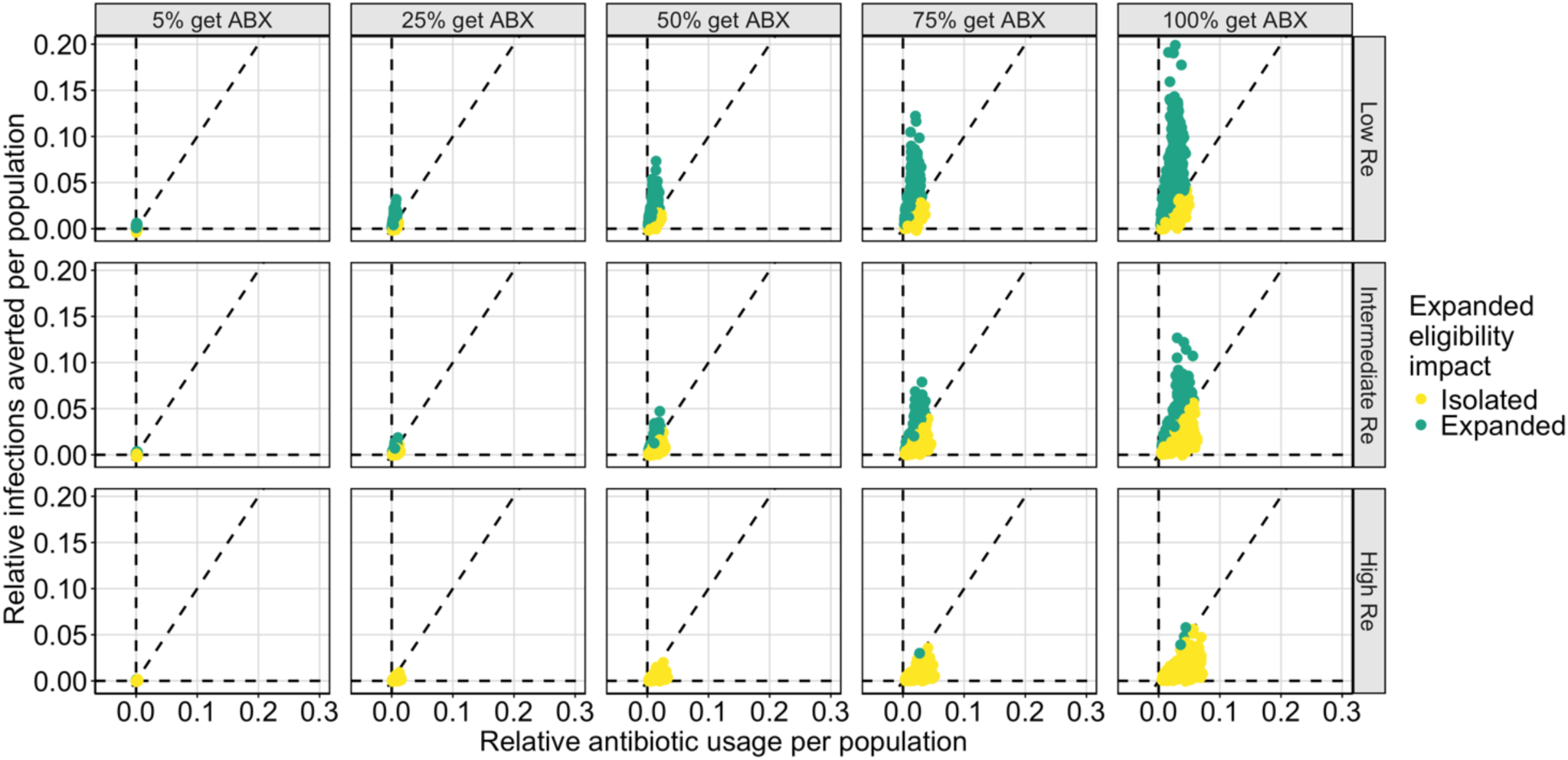
Plot of the population-level impact of expanded antibiotic treatment guidelines with 90%-99% asymptomatic infections. Each plot compares the relative infections averted per population to the relative antibiotic usage per population. The relative reduction in infections is calculated by the ratio of the number of infections in each expanded antibiotic treatment scenario presented compared to the simulation of current antibiotic treatment guidelines (treating no non-severely symptomatic infections with antibiotics) using the same LHS sampled parameters, normalized by population size. Similarly, the relative antibiotic usage is calculated by the ratio of antibiotic doses used in each scenario presented compared to the simulation of current antibiotic treatment guidelines (treating no non-severely symptomatic infections with antibiotics) using the same LHS sampled parameters, normalized by population size. Each plot represents a different proportion of non-severely symptomatic infections seeking care who receive antibiotic treatment (5%, 25%, 50%, 75%, 100%) and a different *R*_*e*_ scenario (low (*R*_*e*_ = 1.3 − 1.5), intermediate (*R*_*e*_ = 1.6 − 2.0), high (*R*_*e*_ = 2.3 − 2.8)). The outcomes are split into three regions by the impact of expanded eligibility criteria: between the dashed line along the x-axis (expanding criteria averts no infections) and the diagonal dashed line (each addition dose of antibiotics deployed averts one infection), yellow points represent simulations in which expanded eligibility only has isolated benefits; between the diagonal dashed line and the vertical dashed line (no additional doses are used to avert infections), green points represent simulations in which expanded eligibility results in each dose preventing more than one additional infection; and in the region left of the vertical dashed line, purple points represent simulations in which expanded eligibility results in fewer doses used over the course of the outbreak than compared to current antibiotic treatment guidelines.

**Figure SI2:**
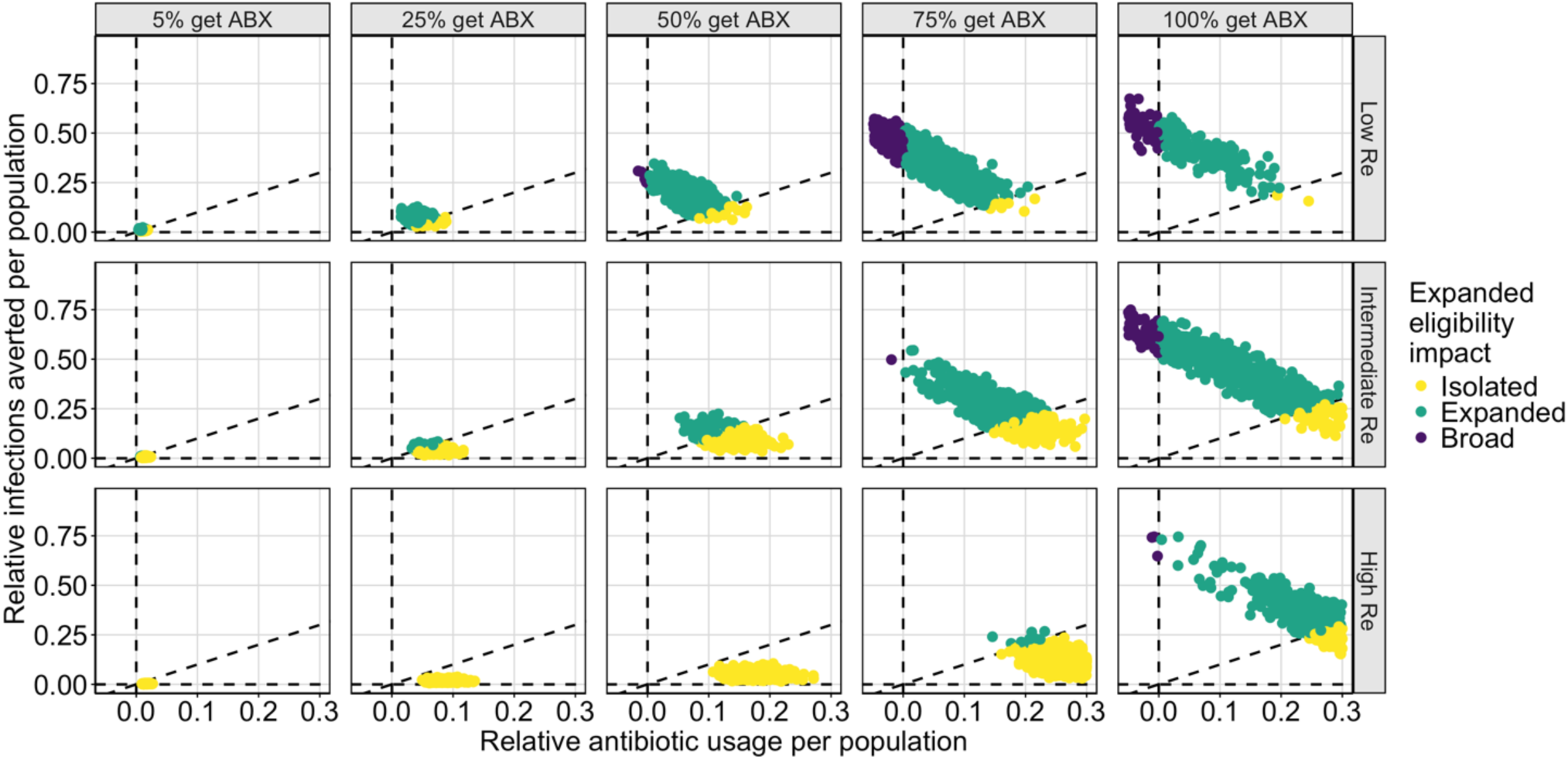
Plot of the population-level impact of expanded antibiotic treatment guidelines with 17%-33% asymptomatic infections. **Figure 3: Plot of the population-level impact of expanded antibiotic treatment guidelines.** Each plot compares the relative infections averted per population to the relative antibiotic usage per population. The relative reduction in infections is calculated by the ratio of the number of infections in each expanded antibiotic treatment scenario presented compared to the simulation of current antibiotic treatment guidelines (treating no non-severely symptomatic infections with antibiotics) using the same LHS sampled parameters, normalized by population size. Similarly, the relative antibiotic usage is calculated by the ratio of antibiotic doses used in each scenario presented compared to the simulation of current antibiotic treatment guidelines (treating no non-severely symptomatic infections with antibiotics) using the same LHS sampled parameters, normalized by population size. Each plot represents a different proportion of non-severely symptomatic infections seeking care who receive antibiotic treatment (5%, 25%, 50%, 75%, 100%) and a different *R*_*e*_scenario (low (*R*_*e*_ =1.3 − 1.5), intermediate (*R*_*e*_ = 1.6 − 2.0), high (*R*_*e*_ = 2.3 − 2.8)). The outcomes are split into three regions by the impact of expanded eligibility criteria: between the dashed line along the x-axis (expanding criteria averts no infections) and the diagonal dashed line (each addition dose of antibiotics deployed averts one infection), yellow points represent simulations in which expanded eligibility only has isolated benefits; between the diagonal dashed line and the vertical dashed line (no additional doses are used to avert infections), green points represent simulations in which expanded eligibility results in each dose preventing more than one additional infection; and in the region left of the vertical dashed line, purple points represent simulations in which expanded eligibility results in fewer doses used over the course of the outbreak than compared to current antibiotic treatment guidelines.

**Figure SI3:**
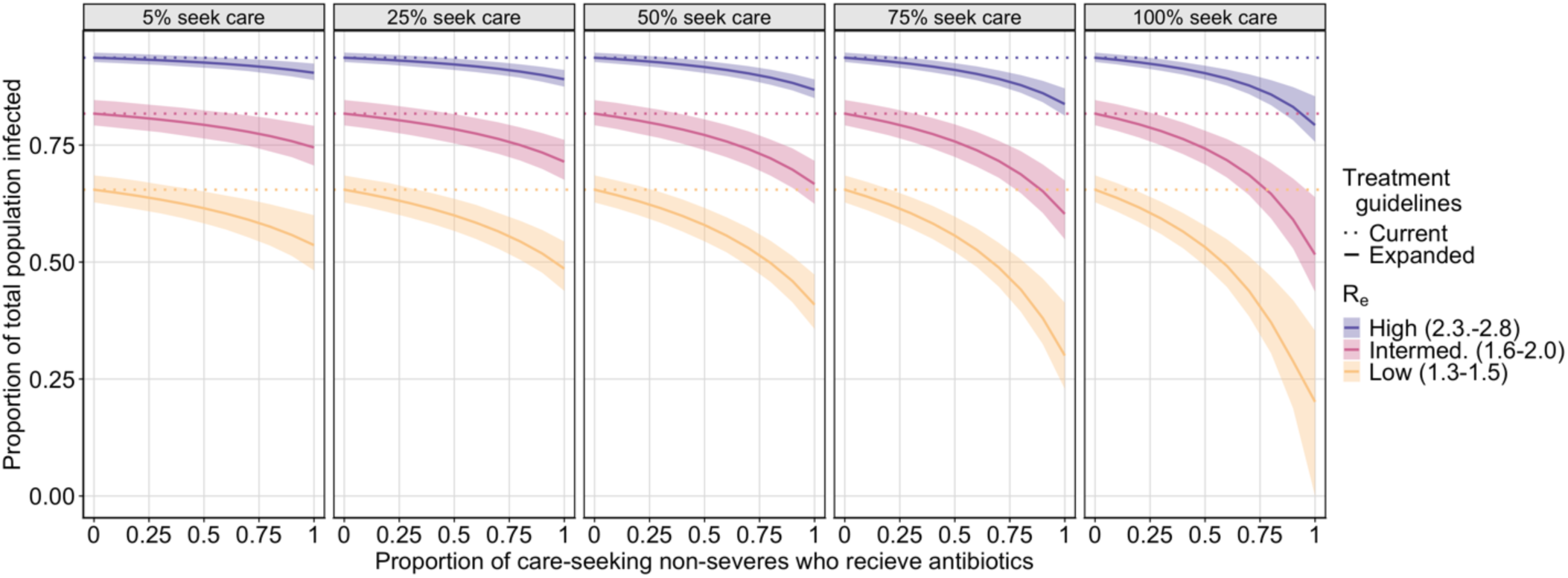
Plot of the final outbreak size by the proportion of care-seeking non-severely symptomatic infections treated with appropriate antibiotics for small proportion asymptomatic (17%-33%). Each plot shows the final proportion of the population infected by the proportion of care-seeking non-severely symptomatic infections who receive appropriate antibiotic treatment for low (yellow), intermediate (pink), and high (purple) effective reproductive numbers, for a different percent of non-severely symptomatic infections who seek care (5%, 25%, 50%, 75%, 100%). The solid line indicates the mean estimate under expanded antibiotic treatment guidelines, the shaded region represents the 25% and 75% quantiles, and the dashed line shows the final size of the outbreak under current antibiotic treatment guidelines (treating no non-severely symptomatic infections).

